# Common Factors in Serious Case Reviews of Child Maltreatment where there is a Medical Cause of Death: Qualitative Thematic Analysis

**DOI:** 10.1101/2021.01.05.20248250

**Authors:** Joanna Garstang, Daisy Eatwell, Peter Sidebotham, Julie Taylor

## Abstract

**Aim:** To identify common factors in Serious Case Reviews (SCRs) where a child has died of a medical cause. Design: Qualitative thematic analysis.

**Background:** SCRs take place when neglect or abuse results in children dying or being seriously harmed. Known key factors within SCRs include parental substance misuse, mental health problems, and domestic abuse. To date there has been no investigation of children who die of a medical cause where there are concerns about child maltreatment.

**Data sources:** A list of SCRs relating to deaths through medical causes was provided from previous coded studies and accessed from the NSPCC National Case Review Repository. Twenty-three SCRs with a medical cause of death from 1^st^ April 2009-31^st^ March 2017 were sourced.

**Results:** Twenty children died of an acute condition and 12 of a chronic condition; 20 of the deaths were unexpected and maltreatment contributed to the deaths of 18 children. Most children were either under one or over 16 at time of death. Many parents were caring for a child with additional vulnerabilities including behavioural issues (6/23), learning difficulties (6/23), mental health issues (5/23) or a chronic medical condition (12/23). Common parental experiences included: domestic violence/abuse (13/23), drug/alcohol misuse (10/23), mental ill health or struggling to cope (7/23), criminal history (11/23), and caring for another vulnerable individual (8/23). Most children lived in a chaotic household characterised by missed medical appointments (18/23), poor school attendance (11/23), poor physical home environment (7/23) and disguised compliance (12/23). All 23 SCRs reported elements of abusive or neglectful parenting. In most there was evidence of cumulative harm, where multiple factors contributed to their premature death. At the time of death 11 children were receiving social care support.

**Conclusion:** While the underlying medical cause of the child’s death was often incurable, the maltreatment that often exacerbated the medical issue could have been prevented.

**Article Summary. Strengths and weaknesses of the study:**

- No other study has analysed SCRs in which children have died of medical causes.
- The most complete dataset possible was used to conduct the robust analysis: SCRs were sourced from the complete list from the Department for Education used for previous national analyses of SCRs.
- Randomly selected SCRs were re-coded by two further researchers to check for any discrepancies in coding, increasing the reliability of results.
- Not all child deaths lead to SCR, even when there are concerns about maltreatment; local areas may differ on their threshold of suspicion; content within SCRs is often variable and inconsistent; so there may be deaths relevant to this study which were not included.
- We only investigated those cases in which a child died, focusing therefore on the worst cases and perhaps missing incidents in which a child had a medical condition and experienced maltreatment but did not die.

## Introduction

Each year around 2000 children in England die aged between the ages of one month and eighteen years. Natural or medical causes of death account for approximately 85%, including deaths from chromosomal, genetic and congenital abnormalities, infections and malignancies, acute and chronic medical conditions (1). Mortality of children in the UK aged one to eighteen years has fallen from 15 to 10 per 100,000 between 2000 and 2018 (2) but UK child mortality remains consistently higher than other countries in Western Europe for many medical conditions (3).

Child maltreatment is the overarching term for when a child has been abused or neglected, including physical, emotional, and sexual abuse (4). One in five adults (18-74 years) report having been a victim of child abuse when under the age of 16 (5) and when neglect or abuse results in serious harm or death of a child (6), the Local Safeguarding Partners (formerly Local Safeguarding Children Boards) undertake a review of the circumstances in order to derive learning to prevent similar situations in the future (4). Previously known as Serious Case Reviews (SCRs), since April 2019 these have been replaced with Child Safeguarding Practice Reviews (7,8), although few of these are yet published and thus analysis continues on SCRs. SCRs only review a small proportion of overall child deaths; in 2017 3% of deaths resulted in an SCR (1).

The links between family backgrounds and childhood neglect or abuse have been heavily researched and are well known within the child protection system (9). Triennial national analyses of SCRs aim to identify key issues, agencies challenges, and inform the government upon the effectiveness of their guidance and further actions needed (10). The most recent triennial review identified recurring factors in SCRs including parental drug and alcohol misuse, criminal behaviour, mental health problems, domestic abuse, and poor engagement with services (11). Different causes of death requiring SCRs have been researched and key themes identified (12). Protective factors that prevent child maltreatment are also well established within the literature including: having a strong mother-child relationship from a young age, positive school environment and (for adolescents) solid peer relationships (13).

Both the safeguarding practice review annual report (14) and the triennial review of SCRs (10) highlight the issue of maltreatment linked to medical causes of death. To date however, we are not aware of any research solely investigating children who die of a medical cause where there are *also* concerns about child abuse or neglect. Such analysis is necessary to identify whether this subgroup have similar background factors to those dying of other causes requiring an SCR (including those deaths directly caused by maltreatment or deaths due to suicide or self-harm), and to provide detailed insight into these factors.

### Aims

This study aimed to identify common factors appearing in SCRs where a child had died of a medical cause. The research questions were:

- What are the common family backgrounds in SCRs where a child has died of a medical cause?
- What are the recurring factors intrinsic to the child in SCRs with a medical cause of death?

## Methods

### Study Design

We used a thematic approach to analysis, based on the qualitative analysis of SCRs undertaken by Garstang and Sidebotham (12). It adhered to the consolidated criteria for reporting qualitative research (COREQ) (15).

As part of the triennial analysis of SCRs, the underlying cause of death was determined for each case. Deaths were determined as being due to medical causes when the primary cause of death was a medical condition regardless of whether abuse or neglect was contributory. This excluded deaths due to inflicted or non-intentional injury, deprivational abuse such as starvation, deaths from external causes, suicide or self-harm, or unexplained causes such as Sudden Unexpected Death in Infancy. A list of SCRs relating to deaths through medical causes was provided from the triennial review process (PS and JG). Published SCRs were accessed from the National Case Review Repository (a collaboration between National Society for the Prevention of Cruelty to Children (NSPCC) and the Association of Independent Local Safeguarding Children Boards)(16) which collates all SCRs in the UK. Only SCRs specifically relating to deaths with medical causes were included. The review covered the last three national analysis periods of 2009-11, 2011-14 and 2014-17 as the SCRs are recent and readily available; these relate to deaths occurring between 1^st^ April 2009-31^st^ March 2017.

### Procedure

Qualitative thematic analysis (17) was used to analyse the SCRs. This method has been effectively used in previous research on similar topics (12). QSR International NVivo V.12 software (18) was used to enable effective analysis and coding of the data. First, all 23 available SCRs were read and summarised, with key ideas for codes being noted. Prior to coding it was decided to use the four domains from the Child Death Review Analysis form (19) as an overarching coding framework. This form is used by the Child Death Overview Panels to analyse factors that may have contributed to a child’s death within four domains: factors intrinsic to the child; social environment; physical environment; and service provision (20). These four domains are linked to those in Bronfenbrenner’s socio-ecological model which explains how both the immediate and more distant environment influences an individual’s development (21). Next, the SCRs were re-read to create initial iterative coding under the four domains. This created many specific codes; these were re-evaluated inductively to create fewer codes to answer the research questions more specifically. These new codes were then revised and refined with second opinions from two experienced researchers. All the SCRs were then re-analysed using the finalised coding system.

Themes were iteratively refined and drawn into multiple theme maps (17) until representative themes and links were found. Finally, to aid in illustration of the subthemes, the quotes from within the code paragraphs were scrutinised to find those which reliably represent the data from within each subtheme.

### Ethics

This study involved analysing published SCRs that are in the public domain. It therefore did not require HRA ethical approval.

### Patient and public involvement

SCRs themselves routinely invite the views of parents and families, although this offer is not always taken up. All are anonymised so we were unable to follow up with individuals. We have a Children and Young People’s Advisory Group whom we intend to involve in dissemination and guidelines for practitioners.

## Results

There were 838 SCRs in the 8 year period, of which 521 related to child deaths. Twenty-six SCRs were identified as relating to deaths from medical causes of which were able to access 23; the remainder were not available on the NSPCC repository. The median age for the index child in the SCR was 8 years (range 2 weeks to 17 years). There were eight girls, eleven boys and in four cases gender was not stated.

Two main themes were identified: predisposing factors and pathway to death. The predisposing factors were of children’s inherent vulnerability which was increased by parenting factors, chaotic homes and professional failings. This then led to a pathway to death (both for children with chronic illness and for previously healthy children) through non-compliance with treatment, failure to seek medical advice, and abuse or neglect. These are illustrated in Figure 1.

**Figure 1:**
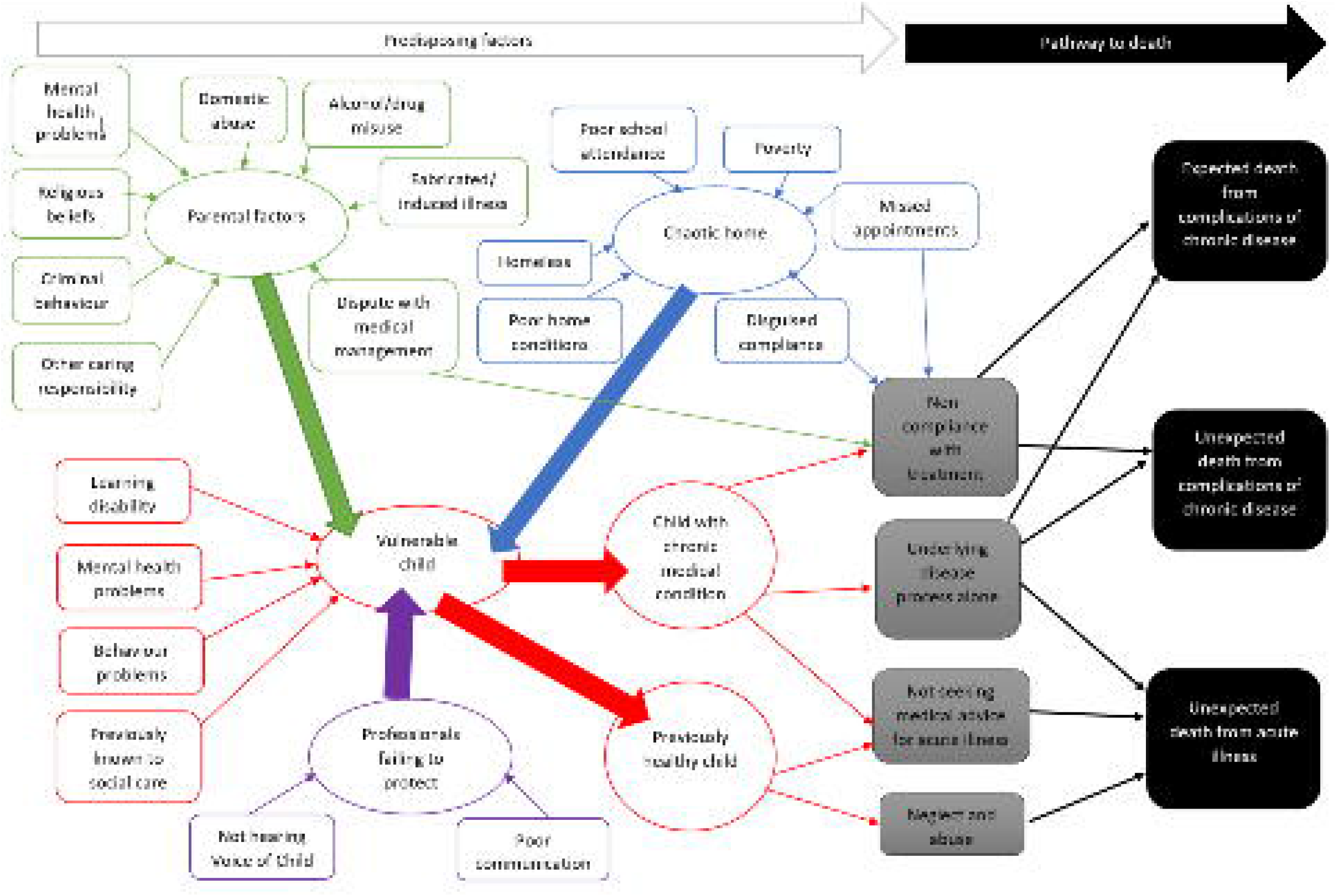
Predisposing factors and pathways to death

### Predisposing factors

#### Vulnerable children

Fourteen children had chronic medical conditions such as asthma, insulin dependent diabetes, cystic fibrosis, morbid obesity or epilepsy which significantly increased their vulnerability. Five children had learning disabilities, six had behavioural problems (e.g. drug misuse, truancy, criminal activities and aggression), and five had mental health problems. Seven children had no additional vulnerabilities but six of these were young infants so inherently vulnerable due to their dependency on parents.

Five children were subject to a child protection plan (CPP) at the time of death, while six had never been referred to social care. In ten families children had previously been subject to CPP, six families had previously been referred to social care, often multiple times, but no further action had resulted from the initial assessment.

#### Parental factors

Domestic violence and abuse (DVA) was a feature in 13 cases; in all cases perpetrators were male and the victim was often abused by multiple different partners over many years. In ten families, parents misused drugs or alcohol while caring for their children; this was a precipitating factor in many DVA episodes. Substance misuse may have directly contributed to the death of a 15-month old child who, although dying of pneumonia, was found to have illicit substances present at post-mortem toxicology. In ten families parents had mental health problems including: depression, suicidal thoughts, panic attacks and borderline personality disorder. DVA, mental health problems and substance misuse co-occurred in five families. Parents in eleven families had criminal records, mainly relating to substance misuse or domestic violence, but three parents had previous convictions for child abuse.

Abuse linked to faith or belief contributed to the death of one child from rickets. The mother followed a strict vegan diet due to her religion, and the infant was exclusively breast-fed, the mother not seeking medical help when the baby became unwell.

There were two cases where parenting beliefs and behaviours in relation to their child’s health needs contributed to the child’s death. In one case, a teenager with features of autism spectrum disorder died as a result of an acute medical condition. In this case, the parents’ views of the child’s needs differed from those of the professionals, and led to the parents removing her from school and support services, and ultimately avoiding hospitalisation when she became acutely unwell. In the other case, the parents’ concerns about the child’s feeding led to force-feeding which contributed to a fatal pneumonia.

Parents in nine families were in dispute with professionals concerning proposed medical treatments, in three cases parents filed multiple complaints about professionals which deflected attention away from the child.

Three families had two children with the same underlying chronic health condition, greatly increasing the strain of caring responsibilities. This was similarly challenging in other families where siblings had behavioural or developmental difficulties.

#### Chaotic household

Chaotic household circumstances contributed to 18 deaths in a variety of different ways. Eight families moved home very frequently, due to evictions, rent arrears and escaping from DVA. This limited access to services; older teenagers were often homeless or moved frequently between family members or hostels and was particularly detrimental to those with chronic health conditions such as diabetes. Poverty was a factor in unstable housing as parents struggled to pay rent and provide for their children. Ten families lived in very poor home conditions, with professionals concerned about the home environment and lack of food. Common issues included: clutter, excrement on floors, unsanitary conditions, rubbish piled up, alcohol bottles, animals and lack of heating. One child with severe asthma was continually exposed to high levels of cigarette smoke despite her carers being informed of the risks this posed.

In 13 families, children or their siblings had poor school attendance which was unrelated to their medical needs. Two children were withdrawn from school as parents disputed the provision available to them, effectively cutting children off from social support and oversight.

In 17 families, children were regularly not brought to medical appointments; this was particularly detrimental for those with chronic medical conditions such as cystic fibrosis, asthma and diabetes as it significantly limited professionals’ ability to offer effective treatment and educate young people and their carers on self-management. Often children were discharged from services due to non-attendance despite their clinical need for treatment, and two families were not registered with GPs.

In 11 families there was evidence of disguised compliance, where parents complied enough to reduce professional concerns only to revert once there was less professional scrutiny. The disguised compliance related to temporary improvements in home conditions, school attendance, medical appointments and treatment regimes.

#### Professionals failing to protect children

Professional failings contributed to children’s vulnerability and poor communication between professionals was apparent in all cases. Within healthcare there was a lack of continuity between professionals with a failure to pass on key information. Discharge summaries of hospital treatment omitted safeguarding concerns such as suspicion that an infant had ingested illicit drugs, or parents delaying seeking medical help for critically ill children. Discharge summaries and outpatient letters were only sent to GPs, excluding Health Visitors and other hospital consultants from key information. Frequently, there seemed to be no overall healthcare professional with a full overview of children’s health needs. Discharge planning meetings were not held despite safeguarding concerns being recognised during hospital admissions. Difficulties with transition to adult services contributed to the deaths of four teenagers, with adult services not recognising the vulnerability of young people who did not comply with medical treatments. One child was not registered with a GP despite having regular appointments in secondary care.

Similarly there was a failure to share information between healthcare and other agencies such as concerns over parental drug and alcohol misuse, mental health issues, and parental refusal to accept medical care for their children. Social care accepted parental accounts of serious disputes with medical staff without seeking further medical information. Strategy meetings were often held for children with complex health needs without any input from medical professionals so other professionals remained unaware of the impact on children of their poor disease management. Within social care there was confusion over the roles of children’s disability teams and assessment teams when safeguarding concerns arose for children with chronic conditions. The different definitions of neglect used by individual agencies limited joint working.

There were issues of poor-quality work in 16 cases: inexperienced social workers had little management supervision, assessments were inadequate, poorly written and confusing, with no mechanism for monitoring parental compliance. Professionals did not reassess children’s vulnerability when situations changed, such as when parents stopped bringing children for appointments. Incidents were often viewed in isolation, not taking into account families’ contexts. Children’s cases were closed and support withdrawn when parents did not engage with Child in Need or Common Assessment Framework plans drawn up by social care. Healthcare professionals did not follow-up concerns around how parents were managing their child’s health needs, and safeguarding was not considered in one child with morbid obesity despite parental refusal to comply with dietary advice.

Twelve SCRs commented that the voice of the child was not heard, with only parents’ views considered even when these were views were potentially harmful such as refusing medical treatment. In many cases the day-to-day experiences of children were unknown to professionals supposedly safeguarding them.

Quotes to illustrate the theme of predisposing factors are shown in table 1.

**Table 1.** Quotes to illustrate the theme of predisposing factors

| Sub-theme | Quote |
| --- | --- |
| Vulnerable children | Parents (the mother and step-father) reported finding the treatment very difficult and noted that his behaviour was also extremely difficult. The child was oppositional (refusing to get washed, dressed, eat, do physiotherapy) and they thought he was low in mood, scratching arms, leaving marks and inducing vomiting when distressed. [Case A] |
| Parental factors | ...it is likely that the young person's mother was presenting the young person's problems as a proxy for her own emotional distress. What this effectively means is that the professionals were trying to 'treat' the mother's emotional, mental health or personality difficulties through the medium of the young person. This was always going to fail, as this is not an effective way to address the mother's needs. [Case B] |
|  | Mother can at times manage some of the practicalities of everyday life with regular support from others, especially her mother. At other times she finds it very hard. Sometimes during Baby's life she needed daily visits from the Family Support Practitioner (FSP) in order to meet the children's basic care needs. [Case C] |
| Chaotic household | Concerns continued relating to home conditions, school attendance and Mother's reliance on teenagers as her main source of support.[Case D] |
| Professionals failing to protect children | A key concern is the omission of the paediatrician in the strategy discussion as it is likely that this would have both occurred face to face at the hospital and provided a comprehensive picture of the risks.... The strategy discussion form is brief on detail and lacks analysis on the rationale for progressing to a Section 47 enquiry, this undermines effective risk assessment. [Case E] |
|  | With the exception of early years practitioners, there was insufficient recognition of day to day experiences and likely long term impact of non life threatening neglect of physical and emotional needs ...The lack of understanding of life in the family from the perspective of the children meant that any analysis of the risk of harm to them was insufficiently informed [Case F] |

### Pathway to death

Children’s deaths were considered as three groups: expected deaths due to complications of chronic disease; unexpected deaths due complications of chronic disease; and unexpected deaths of acute disease. The pathways leading to deaths included non-compliance with treatments for chronic disease, the underlying disease process, parents not seeking medical advice for acute illness, and abuse or neglect. The causes of death for each category are shown in table two.

#### Non-compliance with treatment for chronic disease

Non-compliance with treatment programmes for chronic disease was a causal factor in 10 deaths. Two of these deaths were expected in that the disease had advanced to such an extent that only palliative care was possible, these deaths were from cystic fibrosis and cardiomyopathy due to morbid obesity. The remaining eight deaths were unexpected but could have been avoided with good treatment compliance, this included deaths from acute asthma and diabetic ketoacidosis conditions, from which child deaths are rare.

#### Underlying disease process alone

The underlying acute or chronic illness was severe enough in five cases to lead to death without evidence of non-compliance with treatment, the underlying parental factors, chaotic homes and professional failings contributed to the children’s vulnerability and eventual deaths but was not directly causal. One death was expected, due to a severe underlying genetic condition, four were unexpected due to conditions such as Sudden Unexpected Death in Epilepsy and sepsis.

#### Not seeking medical advice for acute illness

Four deaths were associated with parents not seeking medical advice or disregarding it when their children became ill; these children all died of acute conditions, two had unrelated chronic conditions; causes of death included sepsis and rickets.

#### Abuse and neglect leading to death

In four previously healthy infants, abuse and neglect directly contributed to death although natural disease processes were the underlying cause; this included cases where the final cause of death was pneumonia but there was evidence of non-accidental injury and drug ingestion.

Quotes to illustrate the pathway to death are shown in table 3

**Table 2.**
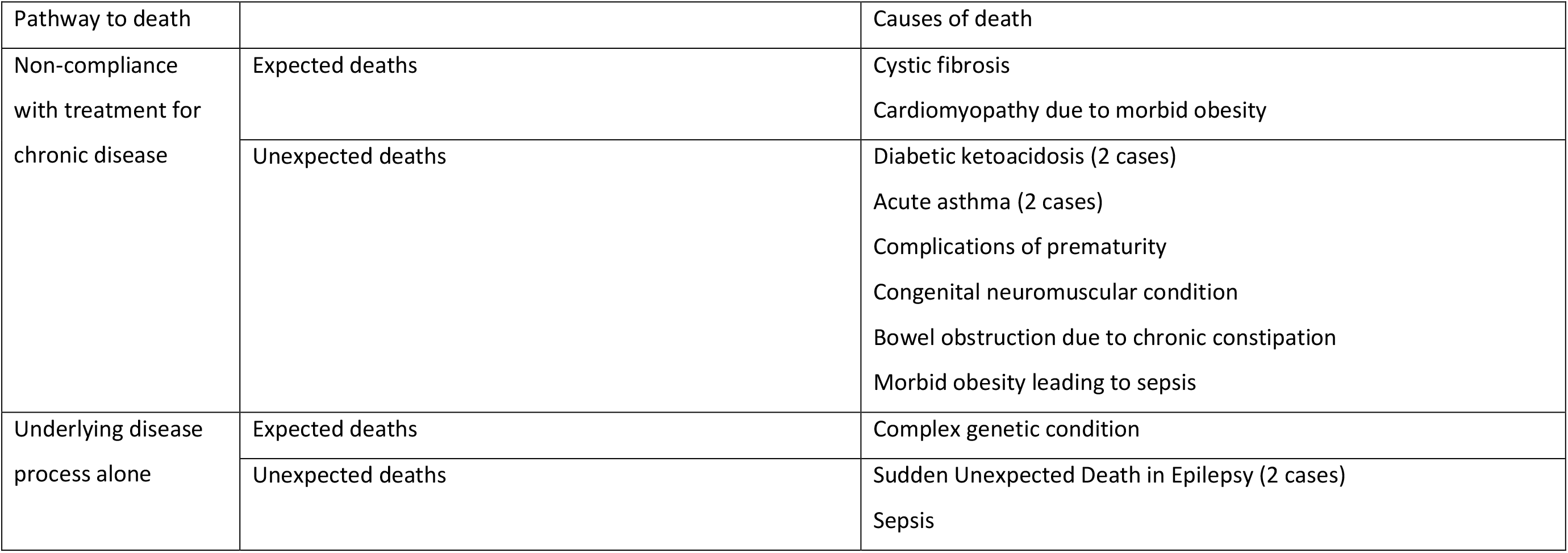
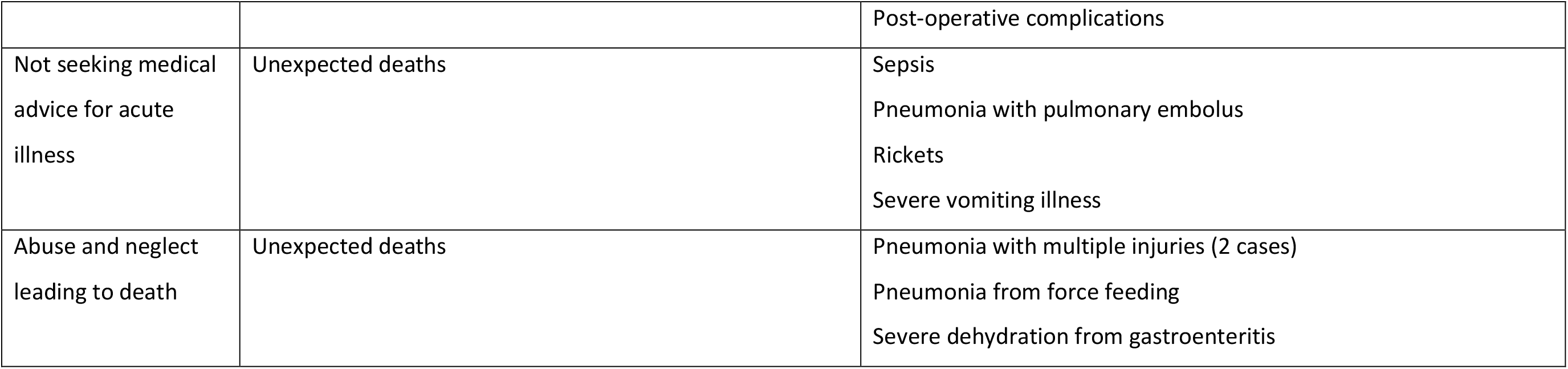
Cause of death

| Pathway to death |  | Causes of death |
| --- | --- | --- |
| Non-compliance with treatment for chronic disease | Expected deaths | Cystic fibrosis<br>Cardiomyopathy due to morbid obesity |
|  | Unexpected deaths | Diabetic ketoacidosis (2 cases)<br>Acute asthma (2 cases)<br>Complications of prematurity<br>Congenital neuromuscular condition<br>Bowel obstruction due to chronic constipation<br>Morbid obesity leading to sepsis |
| Underlying disease process alone | Expected deaths | Complex genetic condition |
|  | Unexpected deaths | Sudden Unexpected Death in Epilepsy (2 cases)<br>Sepsis |
|  |  | Post-operative complications |
| Not seeking medical advice for acute illness | Unexpected deaths | Sepsis<br>Pneumonia with pulmonary embolus<br>Rickets<br>Severe vomiting illness |
| Abuse and neglect leading to death | Unexpected deaths | Pneumonia with multiple injuries (2 cases)<br>Pneumonia from force feeding<br>Severe dehydration from gastroenteritis |

**Table 3.**
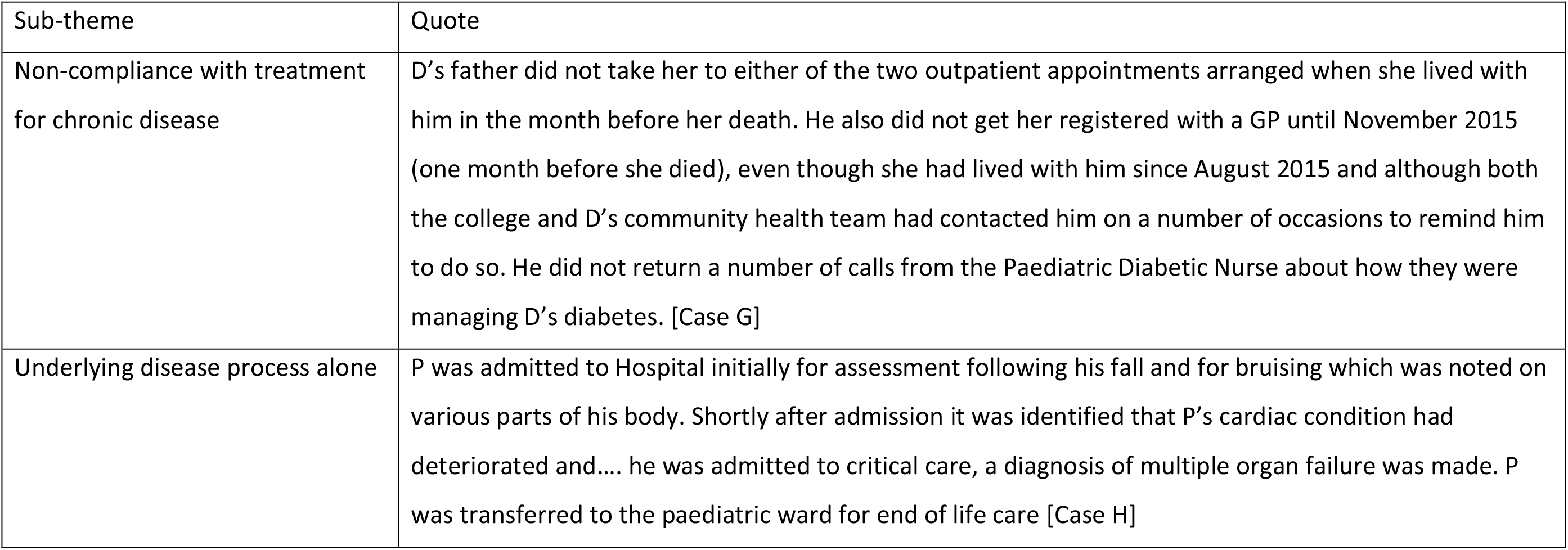
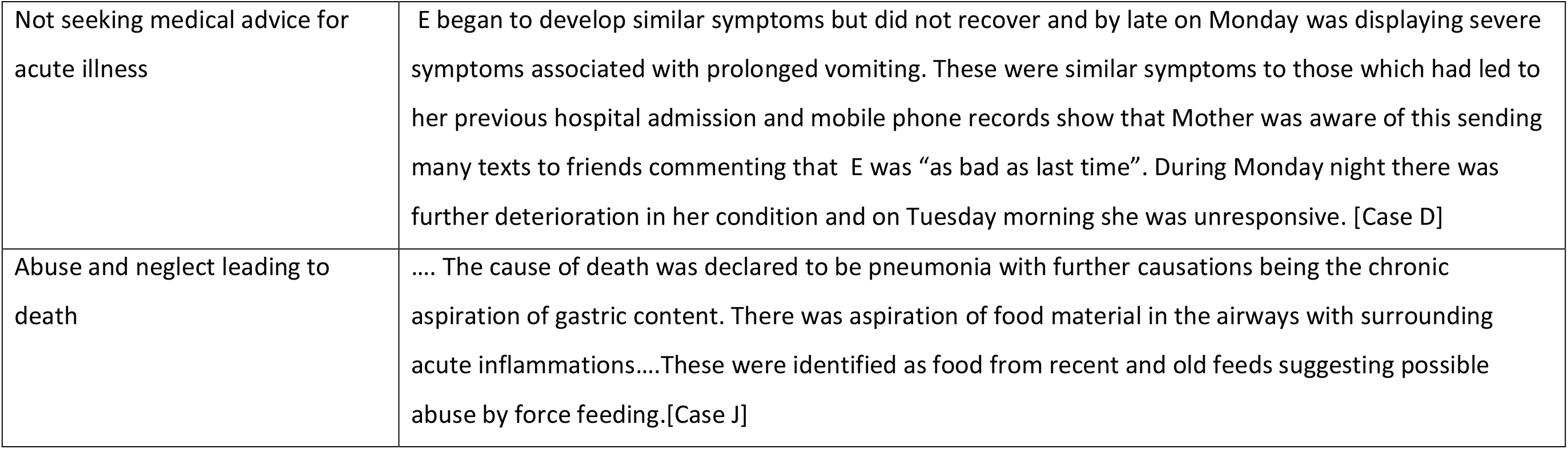
Quotes to illustrate pathway to death

## Discussion

This thematic analysis set out to investigate the common factors within SCRs where there was a medical cause of death. All 23 children were abused or neglected in childhood. Fifteen children had additional vulnerabilities, most commonly a chronic medical condition (12/22), which increased the chaos in the home due to the extra strain in dealing with these vulnerabilities. Chaos was also increased by the parents’ own adversities and these often had a dramatic impact on the children. The chaotic nature of many households caused multiple issues for the children such as missed medical appointments, poor school attendance, housing issues, and abuse or neglect. These recurrent elements resulted in cumulative harm where the combination of many factors increased the child’s vulnerability. Most of the families had child protection service involvement within the child’s lifetime. In 18 of the families the SCRs suggest that abuse or neglect contributed to the child’s death; most died of an acute condition or an acute exacerbation of a chronic condition. Different parental actions may have led to different outcomes in the 20 unexpected deaths.

Many of the key factors found within this study are already well-known within the child protection service (CPS) and established within related literature. In this analysis parental DVA, parental mental health issues, failure to engage with professionals, disguised compliance, poor-quality housing, and parental drug/alcohol misuse were all key factors. These were also frequent elements in a similar study into unexpected infant deaths (12), confirming a well-established link between certain family circumstances and maltreatment. In this study 45% of parents/carers misused drugs/alcohol compared to 66.6% in Garstang and Sidebotham’s analysis (12). Similarly, the triennial analysis of SCRs 2014-17 (10) found that within those SCRs categorised as neglect, 39% contained issues with parental alcohol and drug misuse (though due to differences in the classification of neglect versus all maltreatment, a direct comparison cannot be made). Parenting often becomes a challenge for substance misusing parents and may become neglectful and inconsistent; this may include stricter parenting, emotional withdrawal, increased irritability, and aggressive tendencies towards the child (22).

DVA was a key feature and is a widely recognised factor, for example in the recent annual review of 538 rapid reviews from July 2018-December 2019, 35% of children had DVA as a feature in their life (14). The prevalence of DVA is significantly higher in families in which maltreatment occurs than for the general population of England and Wales (23). DVA constitutes child maltreatment as it affects the child through physical violence and emotional harm (12). Our review has further evidenced the strong link between parental DVA and child maltreatment, but adds new evidence: the negative impact that parental DVA has on children experiencing chronic ill health, potentially contributing to their early death. More must be done to investigate whether children in the home are being abused or neglected and what their experiences are when DVA becomes known to services (24). While there is widespread professional awareness of the risks of neglect, emotional and physical abuse to children because of parental DVA, our research suggests that professionals must further consider the impact of DVA on a child with an illness/disability and ensure adequate cross-agency information sharing. This should include reflecting upon the emotional impact of the DVA on the victim as this may impair their ability to care for the additional needs of their child.

Our study highlights 18 children who died of a medical condition and were not brought to routine or specialist medical appointments. In an analysis of SCRs from 2005-2007, 35% of children likewise had missed appointments (25), and in another study failure to follow up non-attendance at appointments was associated with the child’s death (26). Policies to follow up on children not brought to appointments exist, but more needs to be done to encourage attendance and curiosity into non-attendance must be a priority for all health and social care professionals. Hospital records indicate that those from a deprived background (especially those with parents unable to get time off work) and those with a child protection concern are less likely to attend medical appointments (27). GPs also identify language barriers, cultural differences and, in individuals with complex health needs, unclear letters, as barriers for poor GP attendance (27). Professionals should be aware of the barriers of attendance for their patients, as devising personal solutions may increase attendance.

A major issue within the safeguarding system is premature cessation or stepping down of support. In multiple cases the child’s support from social services was concluded some time before their death often due to non-engagement by parents. In hindsight ongoing provision of support may have prevented the maltreatment or mitigated the effects of ongoing low levels of neglect. Failure to engage with a service is not an appropriate reason to reduce the support for a family, because non-engagement is an indicator of neglect (10). The lack of engagement should indicate the family’s need for further support rather than stepping down support, hence putting the child at increased risk.

## Conclusion

This thematic analysis provides evidence of how children’s additional vulnerabilities, parent’s/carer’s experiences and chaotic households can cumulatively factor into the death of a child with a medical cause. While the underlying medical cause of the child’s death was often incurable, the maltreatment that frequently exacerbated the medical issue could have been prevented. Health practitioners need to be aware of the background family and social factors that may contribute to harm in children with acute or chronic medical needs; likewise, those involved in child protection services need to consider the health needs of children as part of any child protection plan. Further research is needed into how to combat the common issues identified, including non-attendance, parental alcohol and substance abuse, parental DVA, and incorrect discharge from the child protection system.

## Data Availability

Serious case reviews are available at the National Repository

https://learning.nspcc.org.uk/case-reviews/national-case-review-repository

## Author Contributions

JT and JG conceived the idea. JG led the study. DE undertook the initial searches and analysis as part of her undergraduate dissertation. JT and JG supervised. JG, JT and PS undertook further analysis across all SCRs. All authors contributed to the manuscript and read and approved the final version.

## Data sharing statement

Data is drawn from clinical child protection reports of individual children and cannot be shared. The disaggregated and anonymised abstraction files may be shared at reasonable request from the first author.

---

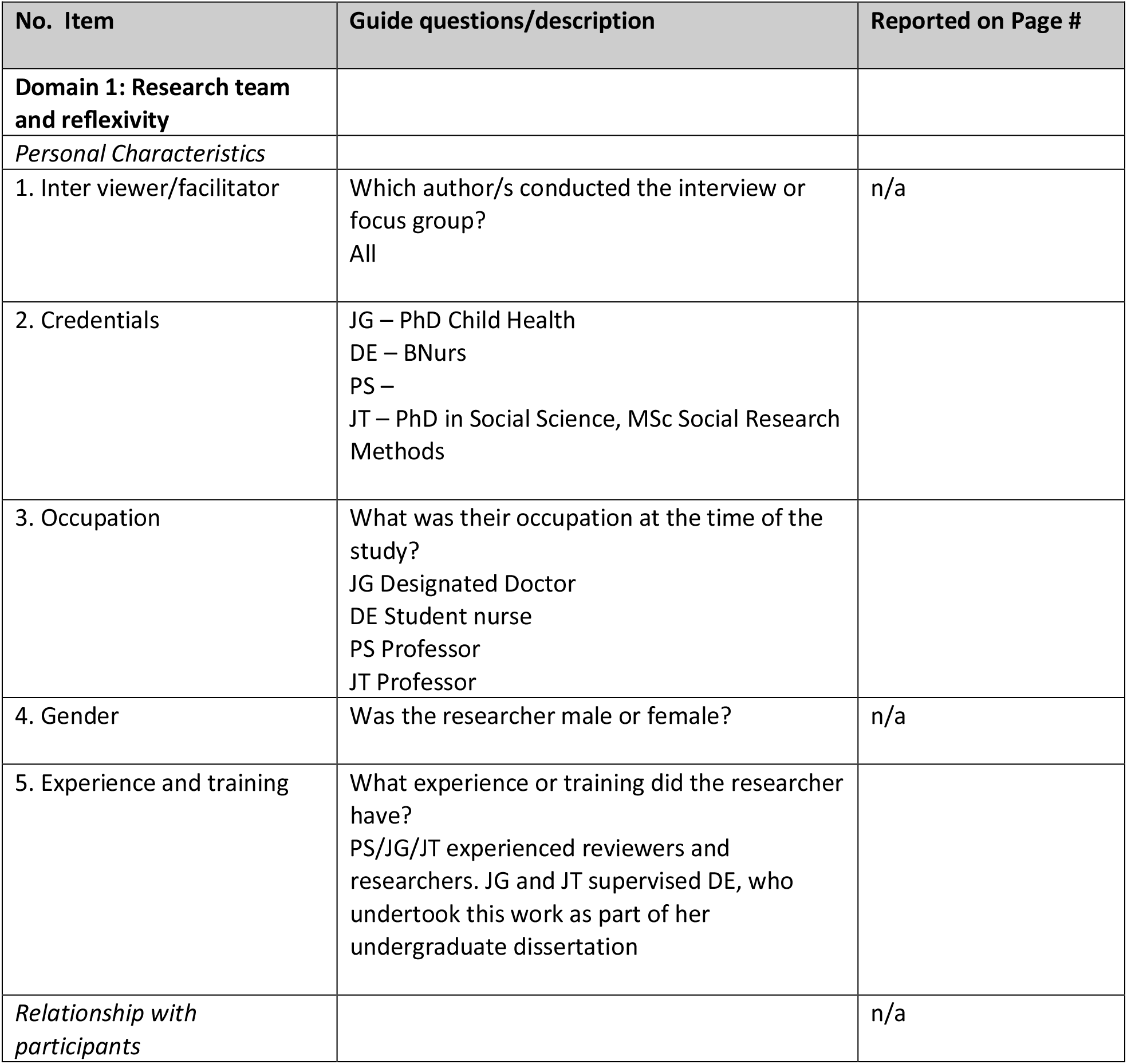
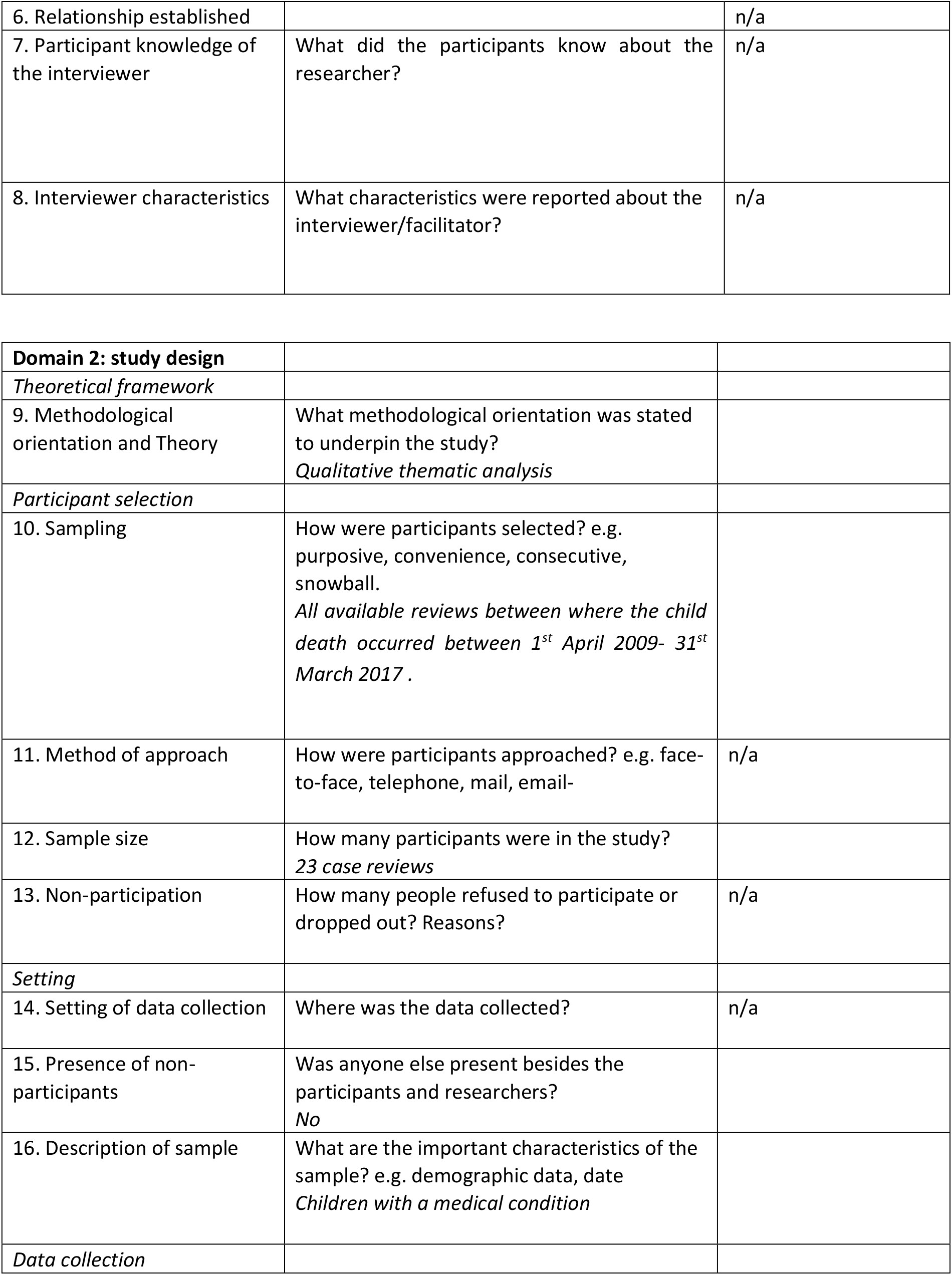
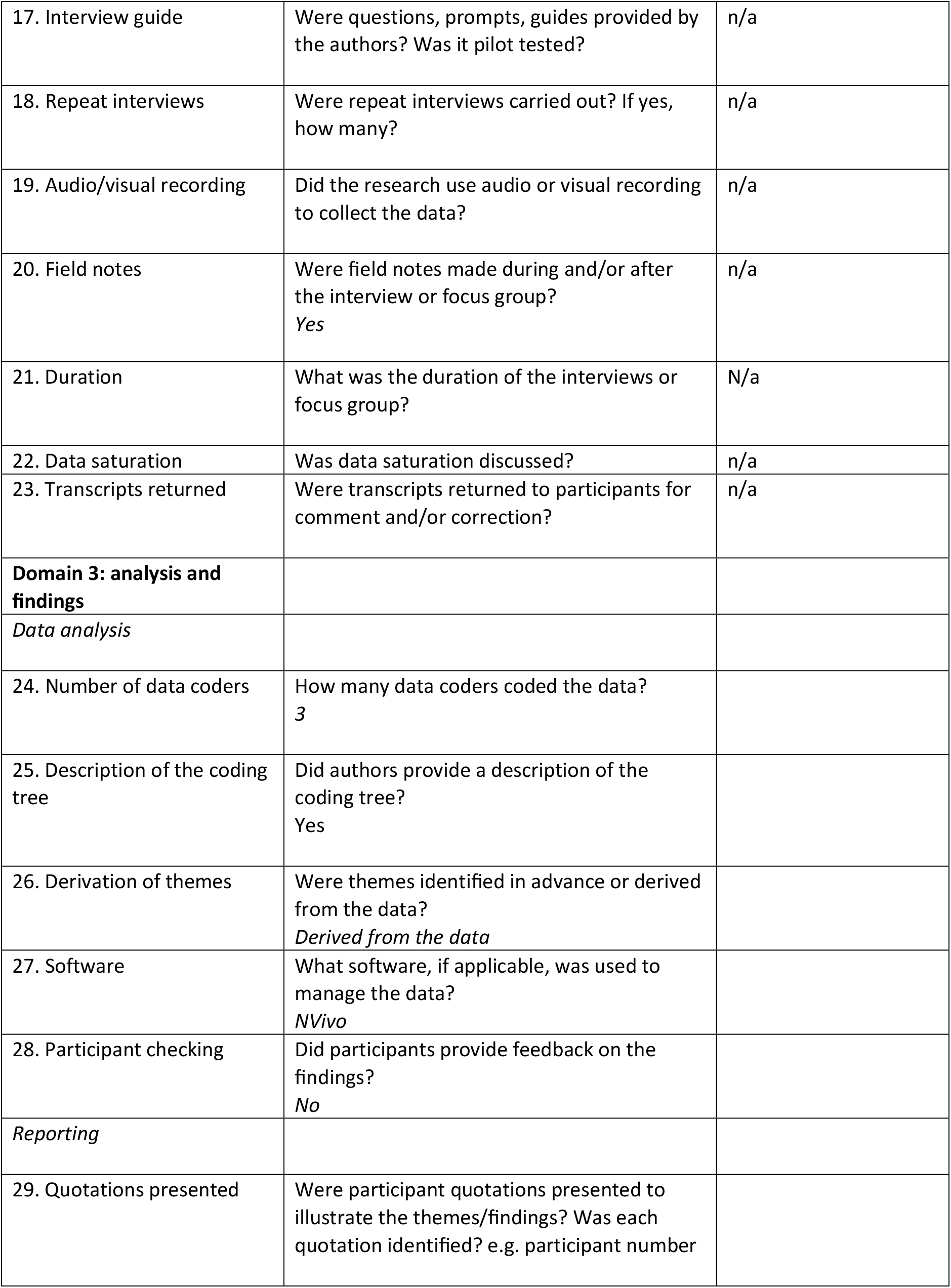
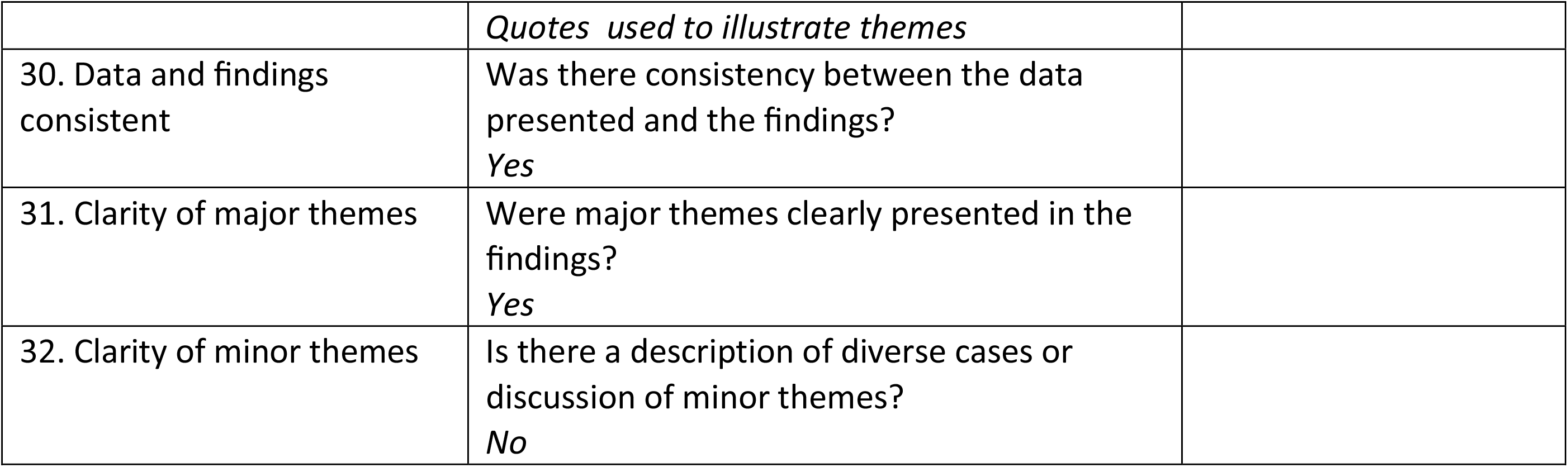

